# Association between use of aspirin use before hospital admission and sepsis outcomes: a population-based cohort study

**DOI:** 10.1101/2020.06.09.20126938

**Authors:** Wan-Ting Hsu, Lorenzo Porta, Tzu-Chun Hsu, Shyr-Chyr Chen, Chien-Chang Lee

**Author notes:** **Correspondence to:** Chien-Chang Lee, MD, ScD, Health Data Science Research Group, National Taiwan University Hospital, Department of Emergency Medicine, National Taiwan University Hospital, No.7, Chung Shan S. Rd., Zhongzheng Dist., Taipei City 100, Taiwan., /. **The two authors contribute equally to this work** (Wan-Ting Hsu, Lorenzo Porta). **Authors’ contributions** W-TH was responsible for statistical analysis, drafting the manuscript, interpretation of the data, and critical revision of the manuscript for important intellectual content. LP was responsible for drafting the manuscript, interpretation of the data and critical revision of the manuscript. T-C.H. preformed data collection, coding, and interpretation. S-CC was involved in grant proposal writing, critical revision of the manuscript for important intellectual content, obtaining funding and supervision. C-CL had full access to all the data in the study and takes responsibility for the integrity of the data and the accuracy of the data analysis, concept and design, critical revision of the manuscript for important intellectual content, obtaining funding and supervision. All authors read and approved the final manuscript.

## Abstract

**Objectives:** Antiplatelet agents have been shown to reduce serum levels of inflammatory markers and may reduce sepsis mortality. A previous randomized clinical trial did not find that aspirin use after hospitalization, compared with placebo, improve acute respiratory distress syndrome within 7 days. We aimed to examine the association between preadmission use of aspirin and sepsis outcome.

**Methods:** We conducted a population-based cohort study based on the National Health Insurance Research Database of Taiwan. The association between aspirin use and 90-day mortality in sepsis patients was determined by Cox proportional hazard models, adjusting for either individual covariates or a propensity score. Restricted mean survival time (RMST) analysis was performed as a sensitivity analysis.

**Results:** Of 52,982 patients with sepsis, 12,776 received preadmission use of aspirin, while 39,081 did not receive any antiplatelets. Use of aspirin before sepsis admission was associated with a decreased risk of 90-day mortality (PS-adjusted HR: 0.89, 95% CI: 0.84-0.93). RMST analysis confirmed the beneficial effect of aspirin which was associated with a 2% increase in survival time (RMST ratio 1.02, 95% CI: 1.01-1.03), when compared to nonuse. On PS adjusted analysis, the odds ratios (OR) of respiratory failure with the preadmission use of aspirin was 0.98 (95% CI: 0.93-1.03), but we did not find any significant association between prior aspirin use and respiratory failure.

**Conclusions:** Our study confirms that prehospital aspirin use was associated with a reduced 90-day mortality rate among sepsis patients, but we did not find any substantial association with respiratory or acute renal failure.

## INTRODUCTION

Sepsis is one of the leading causes of death in hospitalized patients and a growing problem worldwide. In fact, a recent report of the Centers for Disease Control and Prevention (CDC) has shown a 31% increase of sepsis related deaths in the USA from 1999 to 2014^1^. Despite steady improvements in modern medicine, the treatment options for sepsis are limited and, so far, organ support and infection control have remained the two cornerstones of sepsis management^2,3^. However, the global escalation in antimicrobial-resistant bacteria is increasingly compromising the effectiveness of antimicrobial therapy. Thus, treatments directly targeting the host immune response could be the new frontier in sepsis treatment.

Platelets are anucleate cell fragments containing alpha granules, dense granules, and lysosomes, which are activated during infections and systemic inflammations, releasing chemokines, prostaglandins and small molecules and, thus, promoting a pro-inflammatory state and leucocyte migration. Activated platelets have not only been associated with systemic inflammation, but also to thrombotic microangiopathy ^4-6^.

In animal models of endotoxemia and polymicrobial sepsis, platelet ADP inhibitor, such as clopidogrel or monoclonal antibodies directed against the activated platelet glycoprotein IIb/IIIa complex, have been shown to prolong survival^7-9^. Furthermore, in clinical studies on sepsis patients, aspirin and clopidogrel have been shown to reduce serum levels of inflammatory markers, sepsis related multi-organ failure (MOF) and mortality rates ^7-17^. Despite extensive research efforts, previous clinical studies examined the association between sepsis and antiplatelets agent given only after sepsis onset. A randomized clinical trial failed to find a decreased risk of acute respiratory distress syndrome (ARDS) in the post-hospital aspirin use compared with placebo within 7 days^26^. Furthermore, these researches were either performed in ICU populations with limited sample size or in populations of hospitalized patients. A population-based study exploring the effect of use of aspirin before hospital admission on the sepsis outcome has not been published yet. We, therefore, conducted a cohort study to investigate whether preadmission use of aspirin would improve the sepsis outcome. Furthermore, we explored the effect of preadmission use of aspirin on the risk of both septic shock and sepsis-related organ dysfunction.

## METHODS

### Data Source

We performed a population-based cohort study using the NHIRD of Taiwan. Taiwan’s National Health Insurance (NHI) program is a compulsory, government-run insurance plan, covering 99.6% of the island’s population. The NHIRD consisted of one million participants, a systematic multistage sample representative of the age, gender, and regional distribution of the 23 million Taiwanese population in 2001. The included participants were longitudinally followed from 2001 to 2011. The NHIRD contained electronic records on patient demographics, inpatient and outpatient procedures, expenditures, hospital characteristics, prescription details, including brand/generic names of the prescribed drugs, routes of administration, quantities, number of days of supply, and mortality. The NHIRD is particularly suitable for studying the population effect of antiplatelet treatment because aspirin and other antiplatelet medications are not available over the counter in Taiwan. Thus, all prescriptions of antiplatelet agents, alongside dispensing records, are easily retrieved from the NHIRD. The study was approved by the institutional review board of the National Taiwan University Hospital. Informed consents from patients were not required because all the patients were anonymous.

### Patient Population

We conducted a cohort study using the NHIRD. Adult patients (≥ 18 years old) with an emergency department (ED) visit or a hospitalization for sepsis were eligible for inclusion. Sepsis was identified by a code abstraction strategy that combines ICD-9-CM codes for a bacterial or fungal infection and at least one organ/system dysfunctions. This approach, proposed by Angus DC, has been validated extensively in literature ^18^. In fact, not only it is consistent with the latest Sepsis-3 concept, but it also allow the identification of patient populations similar to those identified by a physician review, with a positive predictive value of 71%^19^. We made some modifications by including patients with a single diagnosis of sepsis or septicemia, which was not included in the Angus’s algorithm. In addition, conforming to the Sepsis-3 concept, we defined septic shock by the presence of both diagnostic codes for shock or hypotension and the use of vasopressors. Admission dates of either the ED visit or the hospitalization for sepsis were used as the index date. Subsequently, we followed all patients from the index date until termination of health insurance, death, or the end date of the study period.

### Drug exposure and covariates

Use of aspirin was defined as a record of prescription for at least 7 days in the one-year period prior to the index date. Current use referred to the patients having at least a 7 days prescription length that either covered the index date or had ended within 7 days before the index date. Past use referred to a prescription of aspirin between 91 and 365 days before the index date. Covariate information were collected for each patient on the one-year period before the index sepsis admission. We collected covariate information in the following dimensions: demographics, comorbidities, sources of infection, intensity of healthcare utilization, and co-medications. Detailed covariate information were listed in Table 1.

**Table 1.**
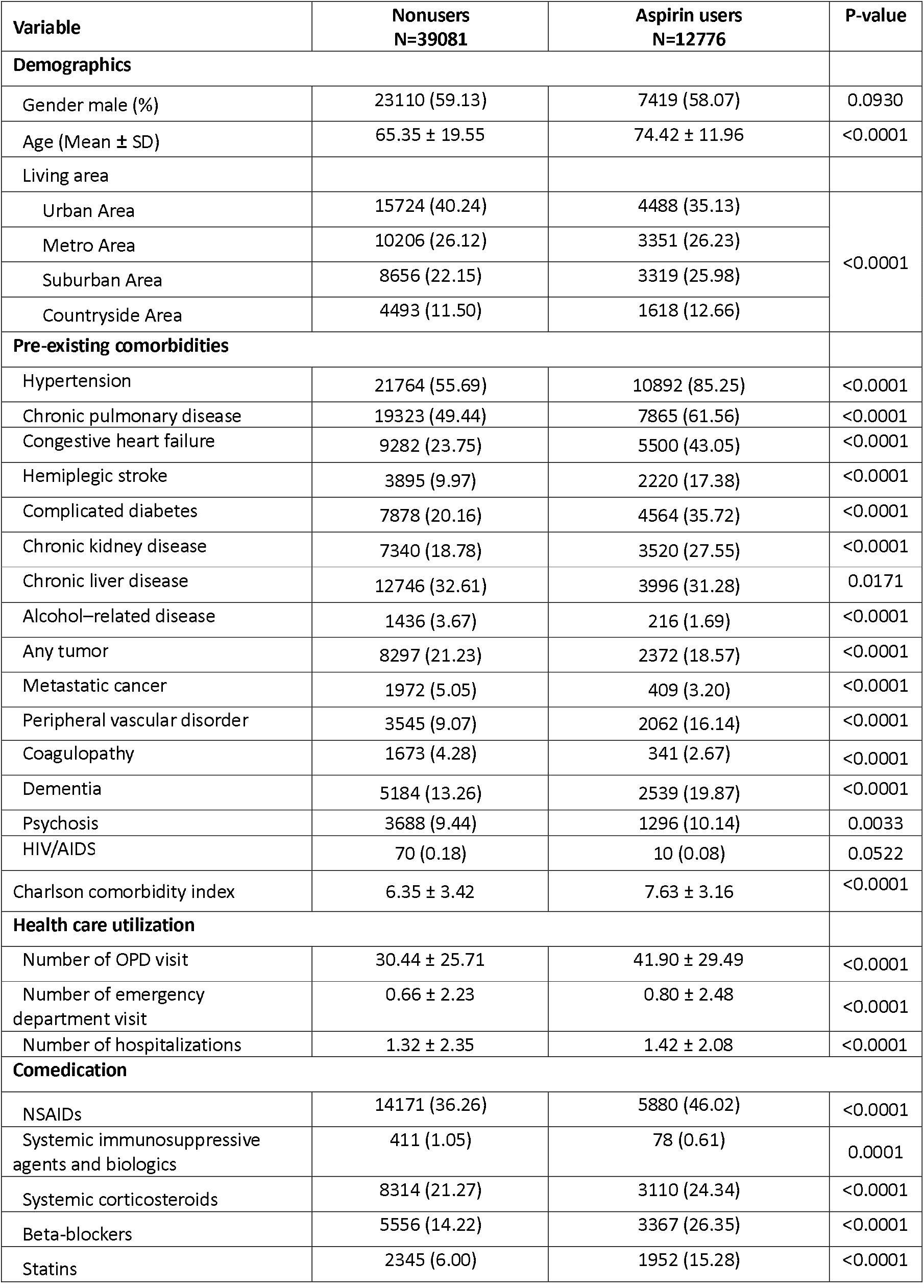
Comparison of baseline characteristics among aspirin users and non-users

| Variable | Nonusers<br>N=39081 | Aspirin users<br>N=12776 | P-value |
| --- | --- | --- | --- |
| **Demographics** |  |  |  |
| Gender male (%) | 23110 (59.13) | 7419 (58.07) | 0.0930 |
| Age (Mean ± SD) | 65.35 ± 19.55 | 74.42 ± 11.96 | <0.0001 |
| Living area |  |  |  |
| Urban Area | 15724 (40.24) | 4488 (35.13) | <0.0001 |
| Metro Area | 10206 (26.12) | 3351 (26.23) |  |
| Suburban Area | 8656 (22.15) | 3319 (25.98) |  |
| Countryside Area | 4493 (11.50) | 1618 (12.66) |  |
| **Pre-existing comorbidities** |  |  |  |
| Hypertension | 21764 (55.69) | 10892 (85.25) | <0.0001 |
| Chronic pulmonary disease | 19323 (49.44) | 7865 (61.56) | <0.0001 |
| Congestive heart failure | 9282 (23.75) | 5500 (43.05) | <0.0001 |
| Hemiplegic stroke | 3895 (9.97) | 2220 (17.38) | <0.0001 |
| Complicated diabetes | 7878 (20.16) | 4564 (35.72) | <0.0001 |
| Chronic kidney disease | 7340 (18.78) | 3520 (27.55) | <0.0001 |
| Chronic liver disease | 12746 (32.61) | 3996 (31.28) | 0.0171 |
| Alcohol-related disease | 1436 (3.67) | 216 (1.69) | <0.0001 |
| Any tumor | 8297 (21.23) | 2372 (18.57) | <0.0001 |
| Metastatic cancer | 1972 (5.05) | 409 (3.20) | <0.0001 |
| Peripheral vascular disorder | 3545 (9.07) | 2062 (16.14) | <0.0001 |
| Coagulopathy | 1673 (4.28) | 341 (2.67) | <0.0001 |
| Dementia | 5184 (13.26) | 2539 (19.87) | <0.0001 |
| Psychosis | 3688 (9.44) | 1296 (10.14) | 0.0033 |
| HIV/AIDS | 70 (0.18) | 10 (0.08) | 0.0522 |
| Charlson comorbidity index | 6.35 ± 3.42 | 7.63 ± 3.16 | <0.0001 |
| **Health care utilization** |  |  |  |
| Number of OPD visit | 30.44 ± 25.71 | 41.90 ± 29.49 | <0.0001 |
| Number of emergency department visit | 0.66 ± 2.23 | 0.80 ± 2.48 | <0.0001 |
| Number of hospitalizations | 1.32 ± 2.35 | 1.42 ± 2.08 | <0.0001 |
| **Comedication** |  |  |  |
| NSAIDs | 14171 (36.26) | 5880 (46.02) | <0.0001 |
| Systemic immunosuppressive agents and biologics | 411 (1.05) | 78 (0.61) | 0.0001 |
| Systemic corticosteroids | 8314 (21.27) | 3110 (24.34) | <0.0001 |
| Beta-blockers | 5556 (14.22) | 3367 (26.35) | <0.0001 |
| Statins | 2345 (6.00) | 1952 (15.28) | <0.0001 |

### Data analysis

Continuous variables were presented with means and standard deviations, while categorical variables were presented with frequencies and percentages. For univariate comparison, we used chi-square tests for categorical variables and analysis of variance for continuous measures. The primary endpoint was 90-day mortality, while the secondary endpoints were septic shock, acute respiratory failure and acute renal failure.

### Propensity score

The propensity score refers to a scalar summary of all observed confounders. Based on the literature review^20^, variables related to aspirin prescription were included in the PS model (Table 1). We plotted the absolute standardized difference graph to show the balance of each covariate after PS matching. The standardized difference after PS matching was less than 10% for each covariate (Supplementary eFigure 1).

### Analysis of association

Univariate association between the use of antiplatelet agents and mortality was evaluated by univariate Cox proportional hazard model. Kaplan-Meier method was used to perform the estimates of cumulative hazard rate for 90-day mortality. The Kaplan-Meier survival curves for each PS-matched cohort pair were graphically plotted. To estimate the hazard ratio and 95% confidence intervals (95% CIs) for the association between an event and each exposure, we used Cox regression models, adjusting for propensity score. The proportional hazards assumption of the Cox model was examined using the log minus log survival (LML) plot. To better quantify the survival difference and gain an outcome difference measure with more clear clinical interpretation, we used the newly developed restricted mean survival time (RMST) method for survival analysis ^21-24^. RMST refers to the mean survival time of all patients within the follow-up period, which can be used to calculate the absolute RMST difference and the relative RMST ratio between the comparison cohorts. We calculated the RMST difference and relative RMST ratio with 95% CIs between aspirin users and PS-matched nonusers. All analyses were conducted with SAS 9.4 for Windows (SAS Institute Inc., Cary, NC, USA). Significant associations were interpreted as p < 0.05.

## RESULTS

### Patient enrollment and patient characteristics

We identified a total of 52,982 patients with sepsis from the NHIRD, of which 12,776 had a preadmission use of aspirin, while 39,081 patients did not receive any antiplatelet medications and, thus, were defined as non-users. Detailed patient inclusion and exclusion process was summarized in supplementary eFigure 2. Compared to nonusers, aspirin users were older, had a higher burden of pre-existing comorbidities, used more healthcare resources and co-medications. The comparison of patient characteristics across aspirin users and non-users was summarized in Table 1.

### Association between antiplatelet use and sepsis mortality

Table 2 presents the association between preadmission use of aspirin and 90-day all-cause mortality. In the crude analysis, aspirin users had an increased risk of 90-day mortality. After adjustment for propensity score, the use of aspirin was associated with a significant decrease in the 90-day mortality risk. Current use of aspirin was associated with a stronger beneficial effect (aHR: 0.89, 95%CI: 0.84-0.93) than past use (aHR: 0.92, 95%CI: 0.88-0.96). To visualize the survival impact of preadmission aspirin treatment on sepsis outcome, we plotted the change of the relative risk over time and the Kaplan-Meier survival curve (Figure 1). The aspirin-associated relative benefit on sepsis mortality was constant over the 90-day following period. Patients taking aspirin prior to the hospital admission were associated with an improved 90-day survival rate (log rank P = 0.0047).

**Figure 1.**
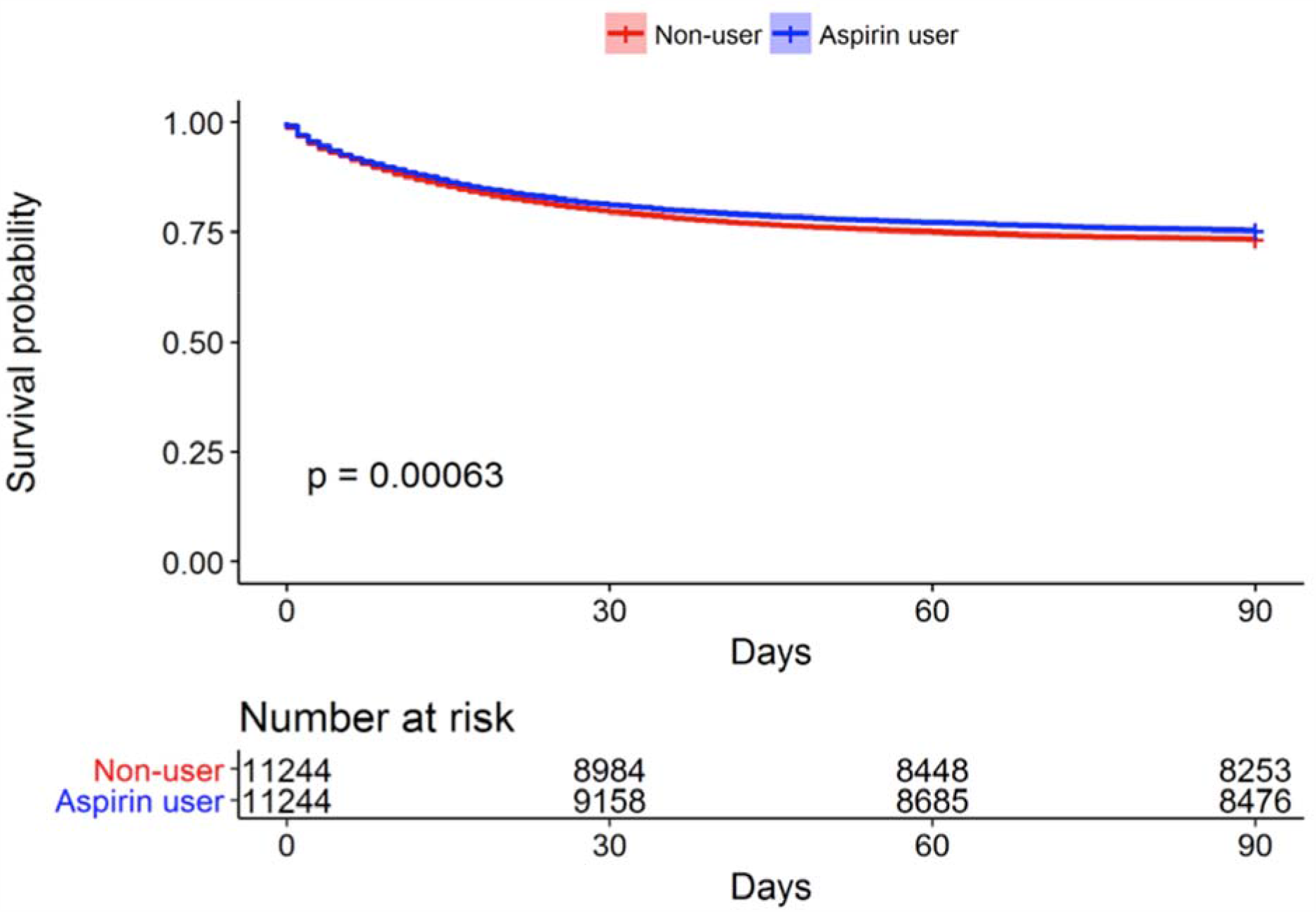
Kaplan-Meier survival curves comparing aspirin users to nonusers in the PS-matched cohort

**Table 2.**
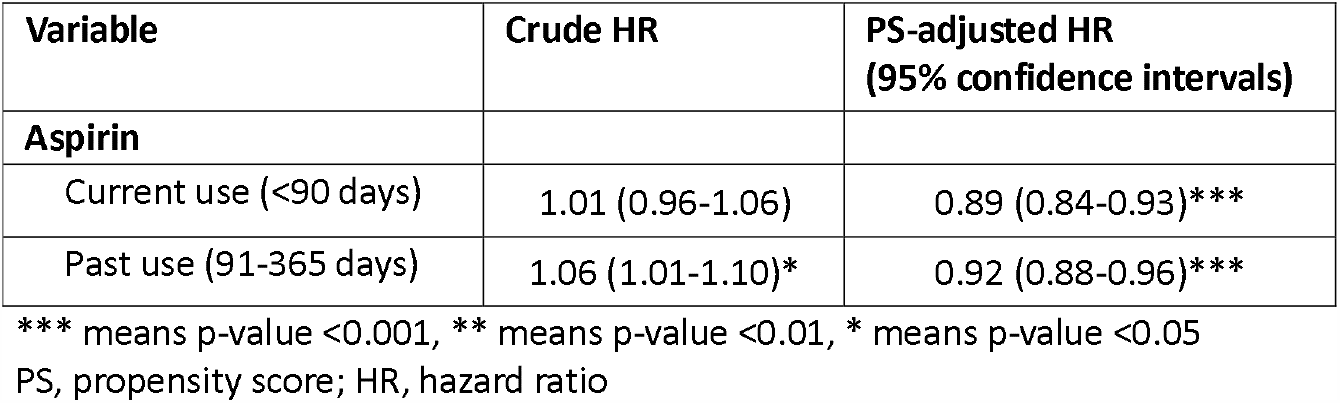
Crude and adjusted effect measure for the association between use of aspirin and risk of sepsis mortality

| Variable | Crude HR | PS-adjusted HR<br>(95% confidence intervals) |
| --- | --- | --- |
| **Aspirin** |  |  |
| Current use (<90 days) | 1.01 (0.96-1.06) | 0.89 (0.84-0.93)*** |
| Past use (91-365 days) | 1.06 (1.01-1.10)* | 0.92 (0.88-0.96)*** |

\*\*\* means p-value <0.001, \*\* means p-value <0.01, \* means p-value <0.05
PS, propensity score; HR, hazard ratio

To verify the robustness of our results and gain a clinically interpretable outcome, we compared the RMST between aspirin users and nonusers in the PS-matched cohorts (Table 3). On RMST analysis, we found that current use of aspirin was associated with an increased mean survival time by 1.31 days (95% CI: 0.42-2.19), when compared to nonuse. Past use of aspirin showed a similar, but attenuated effect (RMST difference 0.90, 95% CI: 0.06-1.75). On the RMST ratio scale, current use of aspirin was associated with a 2% increase in survival time as compared to nonuse (RMST ratio 1.02, 95% CI: 1.01-1.03), while past use was associated with 1% survival time increase (RMST ratio 1.01, 95% CI: 1.00-1.03). Mean survival time for aspirin users within 90 days of hospital discharge was 72.87 days, as compared to 71.56 days for nonusers.

**Table 3.** Restricted mean survival time (RMST) analysis.

| Variable | PS-matched RMST ratio<br>(95% confidence intervals) | PS-matched RMST difference<br>(95% confidence intervals) |
| --- | --- | --- |
| **Aspirin** |  |  |
| Current use (<90 days) | 1.02 (1.01-1.03)** | 1.31 (0.42-2.19)** days |
| Past use (91-365 days) | 1.01 (1.00-1.03)* | 0.90 (0.06-1.75)* days |

\*\*\* means p-value <0.001, \*\* means p-value <0.01, \* means p-value <0.05
PS, propensity score

### Sepsis-related organ dysfunction

We further investigated whether there was a differential effect of aspirin on different organ systems (Table 4). In crude analysis, aspirin users had significant increased odds of septic shock (OR 1.07, 95%CI: 1.02-1.12), respiratory failure (OR 1.20, 95%CI: 1.14-1.26), and acute renal failure (OR 1.25, 95%CI: 1.18-1.33). We, however, did not find any significant association between preadmission use of aspirin and septic shock (PS-adjusted OR 0.99, 95% CI: 0.94-1.04), respiratory failure (OR 0.98, 95%CI: 0.93-1.03) or acute renal failure (OR 1.02, 95%CI: 0.95-1.08).

**Table 4.** Crude and adjusted effect measure for the association between use of aspirin and risk of different endpoints

|  | Crude effect estimate OR<br>(95% confidence interval) | PS adjusted OR<br>(95% confidence interval) |
| --- | --- | --- |
| Septic shock, N= 15,380 | 1.07 (1.02-1.12)* | 0.99 (0.94-1.04) |
| Respiratory failure, N=34,328 | 1.20 (1.14-1.26)*** | 0.98 (0.93-1.03) |
| Acute renal failure, N=8,491 | 1.25 (1.18-1.33)*** | 1.02 (0.95-1.08) |

\*\*\* means p-value <0.001, \*\* means p-value <0.01, \* means p-value <0.05
PS, propensity score; OR, odds ratio

## DISCUSSION

In this national population-based cohort study, we confirmed that the preadmission use of aspirin was associated with improved 90-day survival rates. In fact, current use of aspirin was associated with an 11% reduction in sepsis 30-day mortality. However, the aforementioned aspirin related increase in the survival of sepsis patients was not related to a reduction in specific organ dysfunctions, such as acute respiratory or renal failure. Thus, this beneficial effect of aspirin might be due to other mechanisms other than organ protection

Our results are consistent with the previous published studies, which have demonstrated that pretreatment with low-dose aspirin was associated with better outcomes in patients with community-acquired pneumonia, *Staphylococcus aureus* bloodstream infection, ARDS or any critical illness ^10,13,15,25^. Nonetheless, a multicenter randomized clinical trial did not find that aspirin administration, compared with placebo, significantly reduce the incidence of ARDS at 7 days. We found similar results in our Kaplan-Meier survival curve, which allowed us to visualize the survival analysis of preadmission aspirin use on sepsis outcome^26^. There was no different between non-users and aspirin users within 7 of the 90 day-study period.

Despite the paucity of evidence in literature on the effect of antiplatelet agents on severe infections, there are several plausible mechanisms supporting the observed beneficial effects of aspirin on sepsis patients. At low doses, aspirin inhibits cyclooxygenase-1 (COX-1), causing the inhibition of arachidonic acid, and subsequently of platelet aggregation. At higher doses, aspirin inhibits COX-2 and NF-κB pathway, both of which appear to play key roles in the resolution of inflammation. Since platelets are immune cells that play a role in the activation, amplification and regulation of the inflammation, inhibiting platelets may be beneficial in severe systemic inflammatory responses ^21,27^. Finally, aspirin exerts direct antimicrobial activities. In fact, in a rabbit model of *Staphylococcus aureus endocarditis*, acetylsalicylic acid (ASA) given before and during antimicrobial therapy reduced the vegetation weight significantly ^28^. In another rat model of endocarditis, the combined use of aspirin and ticlopidine was found to be beneficial to infective endocarditis caused by *Enterococcus faecalis* and *Streptococcus gallolyticus* ^29^. In-vitro studies showed that acetylsalicylic acid, the major metabolite of aspirin, activated a stress response regulon sigma fact β, and consequently reduced the virulence of *S. aureus* ^30-32^. Platelets, on the other hand, contribute to biofilm formation; therefore, inhibiting platelets by using low-dose ASA might possibly reduce biofilm formation and its resistance to antimicrobials ^33^.

Results of this study should be interpreted in light of both strengths and weaknesses. The strength of the current study is the availability of a large sepsis cohort. To the best of our knowledge, the current study is by far the largest one to study the effect of preadmission use of aspirin on sepsis outcome. A large population-based database not only allows PS matching between users and nonusers with well-balanced covariates, but also analysis of the effect of several drug combinations. The use of RMST analysis is also a strength, as the validity of proportional hazards assumption is uncertain. The RMST analysis does not rely on any specific assumption; thus, we used this analysis as a sensitivity analysis. RMST analysis also allows quantification of the absolute survival difference and grade the magnitude of clinical benefit.

Nevertheless, there are some limitations in this study. First, lifestyle factors, such as body mass index and smoking were unavailable in the database. Therefore, we used some proxy variables, such as diagnoses of morbid obesity, hyperlipidemia, hypertension, ischemic heart disease and COPD, to adjust obesity and smoking. Second, the potential for healthy user bias remained. Healthy user bias describes the phenomenon for which observed benefits of certain medication actually result from the patients’ behaviors or underlying characteristics. Aspirin users may have higher health awareness, use more healthcare resources, and live a healthier lifestyle than nonusers. However, the inclusion of insurance premium categories, a close proxy for socioeconomic status, and frequency of healthcare utilization in the PS might have reduced the confounding related to healthy users. Finally, it should be noted that, even though the retrospective nature of this study could be interpreted as a limitation, this design was chosen in order to obtain strong statistical results in a big cohort of sepsis patients. Further prospective studies should be performed to fully confirm our findings.

## CONCLUSION

In conclusion, this large population-based study with rigorous statistical analysis provides a strong evidence of the positive effects of the preadmission use of aspirin on sepsis mortality. However, no beneficial effect was found between aspirin use and acute respiratory failure and acute renal failure. Further studies in different populations might be needed to confirm our findings.

## Data Availability

Raw data were generated at National Taiwan
University Health Data Research Center. The derived data that support the findings of this study are available from the corresponding author on reasonable request.

## Acknowledgement

We thank Sonia Hernandez-Diaz, MD, DrPH at Department of Epidemiology and Lee-Jen Wei, PhD at Department of Biostatistics, Harvard T.H. Chan School of Public Health, Boston, MA, USA for comments and suggestions on our study design and manuscript, the staff of the Core Labs, the Department of Medical Research, and National Taiwan University Hospital for technical support, Shih-Hao Lee, and Medical Wisdom Consulting Group for technical assistance in statistical analysis.

## Additional material

Supplementary eFigure 1. Standardized differences of covariates before and after propensity score matching for aspirin users vs. nonusers.

Supplementary eFigure 2. Process of inclusion and exclusion of sepsis patients

